# COVID-19 pandemic risk analytics: Data mining with reliability engineering methods for analyzing spreading behavior and comparison with infectious diseases

**DOI:** 10.1101/2020.11.08.20227322

**Authors:** Stefan Bracke, Alicia Puls

## Abstract

In December 2019, the world was confronted with the outbreak of the respiratory disease COVID-19 (Corona). The first infection (confirmed case) was detected in the City Wuhan, Hubei, China. First, it was an epidemic in China, but in the first quarter of 2020, it evolved into a pandemic, which continues to this day. The COVID-19 pandemic with its incredible speed of spread shows the vulnerability of a globalized and networked world. The first months of the pandemic were characterized by heavy burden on health systems. Worldwide, the population of countries was affected with severe restrictions, like educational system shutdown, public traffic system breakdown or a comprehensive lockdown. The severity of the burden was dependent on many factors, e.g. government, culture or health system. However, the burden happened regarding each country with slight time lags, cf. Bracke et al. (2020). This paper focuses on data analytics regarding infection data of the COVID-19 pandemic. It is a continuation of the research study COVID-19 pandemic data analytics: Data heterogeneity, spreading behavior, and lockdown impact, published by Bracke et al. (2020). The goal of this assessment is the evaluation/analysis of infection data mining considering model uncertainty, pandemic spreading behavior with lockdown impact and early second wave in Germany, Italy, Japan, New Zealand and France. Furthermore, a comparison with other infectious diseases (measles and influenza) is made. The used data base from Johns Hopkins University (JHU) runs from 01/22/2020 until 09/22/2020 with daily data, the dynamic development after 09/22/2020 is not considered. The measles/influenza analytics are based on Robert Koch Institute (RKI) data base 09/22/2020. Statistical models and methods from reliability engineering like Weibull distribution model or trend test are used to analyze the occurrence of infection.

## 1 Introduction

In December 2019, the world was confronted with the outbreak of the respiratory disease COVID-19 (“Corona”). The first infection – confirmed case – was detected in the City Wuhan, Hubei, China. First, it was an epidemic in China, but in the first quarter of 2020, it evolved into a pandemic, which continues to this day.

The COVID-19 pandemic with its incredible speed of spread shows the vulnerability of a globalized and networked world. The first months of the pandemic were characterized by heavy burden on health systems. Worldwide, the population of countries was affected with severe restrictions, like educational system shutdown, public traffic system breakdown or a comprehensive lockdown. The severity of the burden was dependent on many factors, e.g. government, culture or health system. However, the burden happened regarding each country with slight time lags, cf. Bracke et al. (2020).

This paper focuses on data analytics regarding infection data of the COVID-19 pandemic. It is a continuation of the research study “COVID-19 pandemic data analytics: Data heterogeneity, spreading behavior, and lockdown impact”, published by Bracke et al. (2020). The goal of this assessment is the evaluation/analysis of infection data mining considering model uncertainty, pandemic spreading behavior with lockdown impact and early second wave in Germany, Italy, Japan, New Zealand and France. Furthermore, a comparison with other infectious diseases – measles and influenza - is made.

The used data base from Johns Hopkins University (JHU) runs from 01/22/2020 until 09/22/2020 with daily data, the dynamic development after 09/22/2020 is not considered. The measles/influenza analytics are based on Robert Koch Institute (RKI) data base 09/22/2020. Statistical models and methods from reliability engineering like Weibull distribution model or trend test are used to analyze the occurrence of infection.

## 2 Goal of Research Study

The overarching goal is the analysis of the development of infection occurrence within the mid-early COVID-19 pandemic time (12.2019 – 09.2020). The detailed topics are as follows:

1. Overview with respect to data quality and the impact on uncertainty,
2. Impact of lockdown based on spreading behavior analytics,
3. Detection and analyzing of spreading behavior in the early second wave,
4. Comparison of COVID-19 spreading behavior with other infectious diseases (influenza/measles).

These topics are discussed based on data from five different reference countries.

The selection of the countries was based on following characteristics:

1. Germany: Data quality/access, lockdown: distance regulations and restrictions on contact,
2. Italy: First massive outbreak in Europe, hard lockdown,
3. Japan: Soft lockdown, socially usual high hygienic standard,
4. New Zealand: hard lockdown, effective border closures and soft second wave,
5. France: hard lockdown in first wave and strong spreading in second wave.

## 3 Methods

This section shows the statistic fundamentals for analyzing the COVID-19 pandemic data. The spreading behavior and the impact of the lockdown regarding different countries is analyzed by using the Weibull distribution model. The Weibull distribution model is frequently used within reliability engineering and risk analytics, cf. Birolini (2017). The applicability of this model for the evaluation of occurrence of infection is based on the exponential progress.

In addition to classical methods of virology such as the SIR model (cf. Kermack and McKendrik (1927)), e.g. applied for COVID-19 in D’Arienzo and Coniglio (2020) in combination with basic reproduction number, the Weibull distribution model offers the possibility, to gain knowledge with regard to the infection development. The easy interpretability of the Weibull parameters allows the analysis of the spreading behavior, in particular the spreading speed. This is the first advantage in comparison to the use of an exponential distribution model. Second advantage is the the normalization of the Weibull distribution function: It allows an easy comparison of measurement data based on different time ranges (samples).

Therefore, the analysis of the spreading behavior before and after lockdown as well as the comparison of COVID-19 with other infectious diseases are based on the determination and interpretation of these Weibull parameters. While the analysis of the second wave is also done with the Weibull model, the detection is conducted with a Cox-Stuart Trend Test (Significance test).

### 3.1 Weibull distribution model

The two-parameter Weibull distribution model is given based on Eq. (1), cf. Weibull (1951).

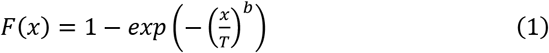

The parameters, besides the term life span variable x, are scale parameter T (in lifetime analysis: characteristic life span) and shape parameter b. By variating parameter b, different failure rates can be described, therefore the Weibull model can be flexibly used for different applications, cf. Rinne (2008). The shape parameter b gives hints regarding the character of the failure period: early failure period, random failure or operation time related failure behavior. The Weibull parameters are estimated by using the Maximum Likelihood Estimator (MLE), cf. Fisher (1912).

In occurrence of infection, the shape parameter b as gradient of the model is interpreted as spreading speed (transfer thinking of reliability engineering: here, shape parameter b describes the occurrence of damage cases within a product fleet). The scale parameter T gives another hint of the spreading speed considering the first infection case. Representing the x value of the probability 0.633%, T indicates while comparing different models (e.g.: countries) how fast the infection cases are progressing in relation to the total days.

### 3.2 Cox-Stuart Trend Test

The Cox Stuart trend test is a non-parametric statistical test for detecting trends in a sample, based on the Binomial distribution. The data is divided in the midpoint into two sequences and the paired difference D is build. For the detection of the second wave, the one-sided form of the test is used to determine an upward trend. Therefore, the number of the positive signs in D is defined as S+. The null hypothesis states that S+ follows a binomial distribution with the number of experiments n as number of elements of D and a probability 0.5. If the p-value of the test is smaller than the significance level alpha, the null hypothesis is rejected and an up-trend is confirmed; cf. Cox and Stuart (1955), Papula (2017):

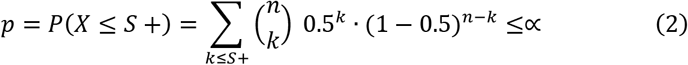

For detecting the second wave, the confirmed daily cases are analyzed as time series, so the Cox Stuart trend test is applicable.

## 4 Data Base

The base of operations for the presented research study are the documentation of the worldwide infection data of the Johns Hopkins University (JHU). The COVID-19 dashboard by the Center for Systems Science and Engineering (CSSE) at Johns Hopkins University (2020) documented confirmed cases, recovered cases as well as death cases regarding countries and regions, starting at 01-22-2020, cf. JHU (2020). Furthermore, the data was compared with the Robert Koch Institute (RKI) on a case-by-case basis; cf. RKI (2020, September).

As a frame for the data basis for the presented study, Table 1 shows an overview of key data with regard to 1^st^ infection (confirmed case) and lockdown by countries, which are in focus in this paper. For the presented research work, the data regarding the first infection and the lockdown are relevant for the analyzed country.

**Table 1:** Dates of first infection and lockdown per country.

| Country | 1st infection | lockdown |
| --- | --- | --- |
| Germany | 01/28/2020 | 03/22/2020 |
| Italy | 01/28/2020 | 03/09/2020 |
| Japan | 01/16/2020 | 03/28/2020 |
| France | 01/24/2020 | 03/17/2020 |
| New Zealand | 02/28/2020 | 03/26/2020 |

Note: The date “1st infection” represents the index case, the first documented case. The design of the lockdown had different characteristics: The Italy lockdown includes the prohibition for leaving home (except for necessities). The lockdown in Japan was a strong recommendation of the government; a real lockdown cannot be enforced considering the Japanese constitution. In Germany, distance regulations come into effect since 22 March. The lockdown in France was characterized by curfew and movement restriction. The lockdown in New Zealand was focusing on strict border closures, favored by island location.

## 5 Data quality and impact on uncertainty

It must be considered, that the data quality within the JHU database is different, caused by reasons with relation to the reported countries: E.g., data can be incomplete and censored, depending on the collection and reporting system of the country. Furthermore, the different definitions of facts (e.g. death case: death with COVID-19 or because of COVID-19) has an impact on the data. This section gives a brief overview of the uncertainty with regard to data acquisition, for detailed explanations cf. Bracke et al. (2020).

First of all the type of measuring method has to be considered. Three aspects can be mentioned:

- Criteria for testing (test strategy, e.g. symptom-based or area-wide),
- Reporting system (reporting procedure),
- Accessibility of health department (e.g. weekend-impact).

The definition of a case of illness (confirmed case) is also relevant for the database. Other relevant definitions are infection, recovered or death case.

The post mortem analysis has a strong impact on death cases and can be different regarding countries and regions (e.g. Post mortem analyses required by law or individual decision by the doctor who determines the cause of death).

Apart from the political measures and differences in measurement and definitions considered here, many other factors influence virus spread and thus the data situation. Some of these uncertainty factors are (without claiming to be conclusive); cf. Dimmock et al. (2016):

- Seasonality and climatic effects, cf. Sajadi et al. (2020),
- Frequency of susceptible individuals in the population, like urbanity and persons in agglomerations (population density),
- Differences in behavior, e.g. cultural or climatic determined,
- Type of treatment, cf. Gattinoni et al. (2020).

With the knowledge that these uncertainties occur, the comparative data is kept as constant as possible. Therefore:

- The data is differentiated between before and after lockdown.
- Comparison of countries with similar industrialization standard.
- Ranked data is used and the time is normalized to the arrival date.

In addition, the results are checked for plausibility and uncertainties during the analyses.

## 6 Analyses of the COVID-19 spreading behavior

This section focusses on COVID-19 data analytics: The analyses of the spreading behavior within different countries in first wave, considering the spreading before and after lockdown; and the analyses of the early second wave.

### 6.1 Infection (confirmed cases) before lockdown

The comparison of the development of the infection in Germany, Italy, Japan, France and New Zealand with the focus on confirmed cases is shown in Fig. 1 (log-log-scale). Weibull distribution models (cf. Eq. 1) are fitted based on the cumulative cases; the Weibull parameters are estimated by using the MLE and shown in Table 2; shape parameter incl. confidence belt with confidence level γ = 0.95. The Weibull model and the parameters shows the infection development in a sound way: The Weibull fit is based on the data range first known infection case (day 1) until the lockdown related to the analyzed country. Not all uncertainty factors could be correctively considered caused by unavailable information. However, the different characteristics of the spreading behavior in the countries under consideration can be seen.

**Table 2:** Weibull model parameters before lockdown starting with 1st infection case (cumulative confirmed cases related to analyzed country). Confidence level γ = 0.95.

| Country | cases | T [d] | Shape b [confidence belt] |
| --- | --- | --- | --- |
| Germany | 22,213 | 53 | $19.61 \leq 19.82 \leq 20.03$ |
| Italy | 9,172 | 38 | $12.91 \leq 13.12 \leq 13.34$ |
| Japan | 1,466 | 52 | $4.60 \leq 4.79 \leq 5.00$ |
| France | 7,652 | 52 | $17.48 \leq 17.80 \leq 18.12$ |
| New Zealand | 283 | 26 | $9.95 \leq 10.96 \leq 12.01$ |

**Fig. 1:**
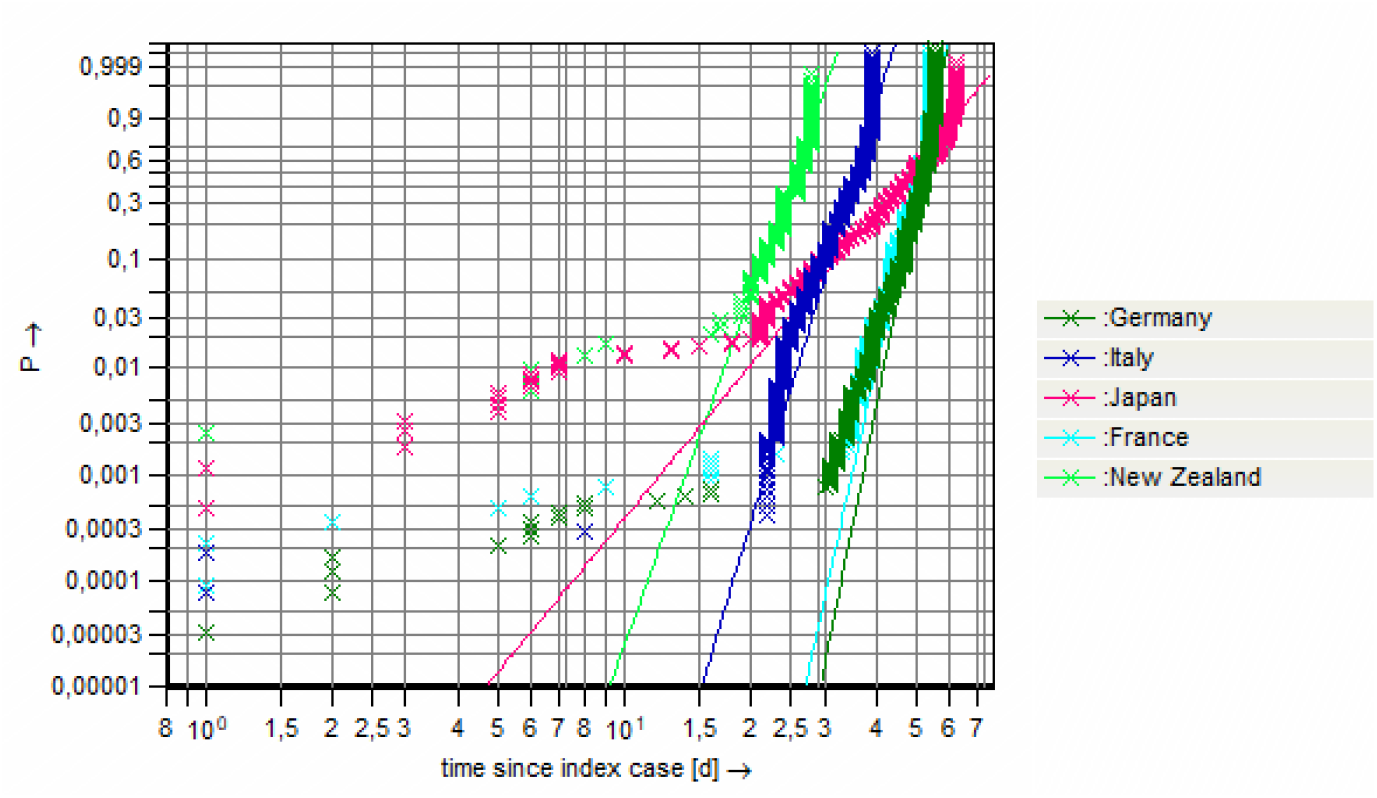
Weibull distribution models, representing the infection development, confirmed cases, interval 1st infection case (index case) until lockdown related to the analyzed countries (log-log-scale).

The Weibull models show the different infection developments: The shape parameter represents the gradient and can be interpreted as the spreading speed of the infection within the population. A steep curve means a high spreading speed.

Japan shows the lowest shape parameter (spreading speed). The possible reason for this effect can be the socially usual high hygienic standard (e.g. wearing masks, social distance), which was additionally enforced by the COVID-19 pandemic. The shape parameters Germany, Italy and France are on a similar level, this gives a clear hint regarding the high spreading speed (behavior) within a short time period (few weeks) in Europe. The small number of cases and the lower T of New Zealand can be explained with the fast decision and implementation of the lockdown (about one month) in comparison to the other analyzed countries (about two months), cf. Table 1.

Note: In reliability engineering, typical wear out mechanism can have 1.5 ≤ b ≤ 3. Brittle failures can have b ∼ 8. Thinking transfer (from reliability engineering point of view): The shape parameters regarding COVID-19 spreading behavior in Germany, Italy, France (cf. Table 2) show a very strong gradient respectively spreading speed within the population in comparison to strong failure spreading behaviors within technical product fleets (e.g. automobiles) in use phase; e.g. cf. Sochacki and Bracke (2017).

### 6.2 Infection (confirmed cases) after lockdown

The measure lockdown causes a significant change regarding the spreading speed (behavior) with respect to the analyzed countries. Fig. 2 shows the Weibull distribution model fits based on the confirmed cases data after lockdown considering a time span of approximately 28 days. The Weibull distribution model describes once again the distribution of the measurement data (confirmed cases) after the lock down. It is not a prediction of the expectable future confirmed cases.

**Fig. 2:**
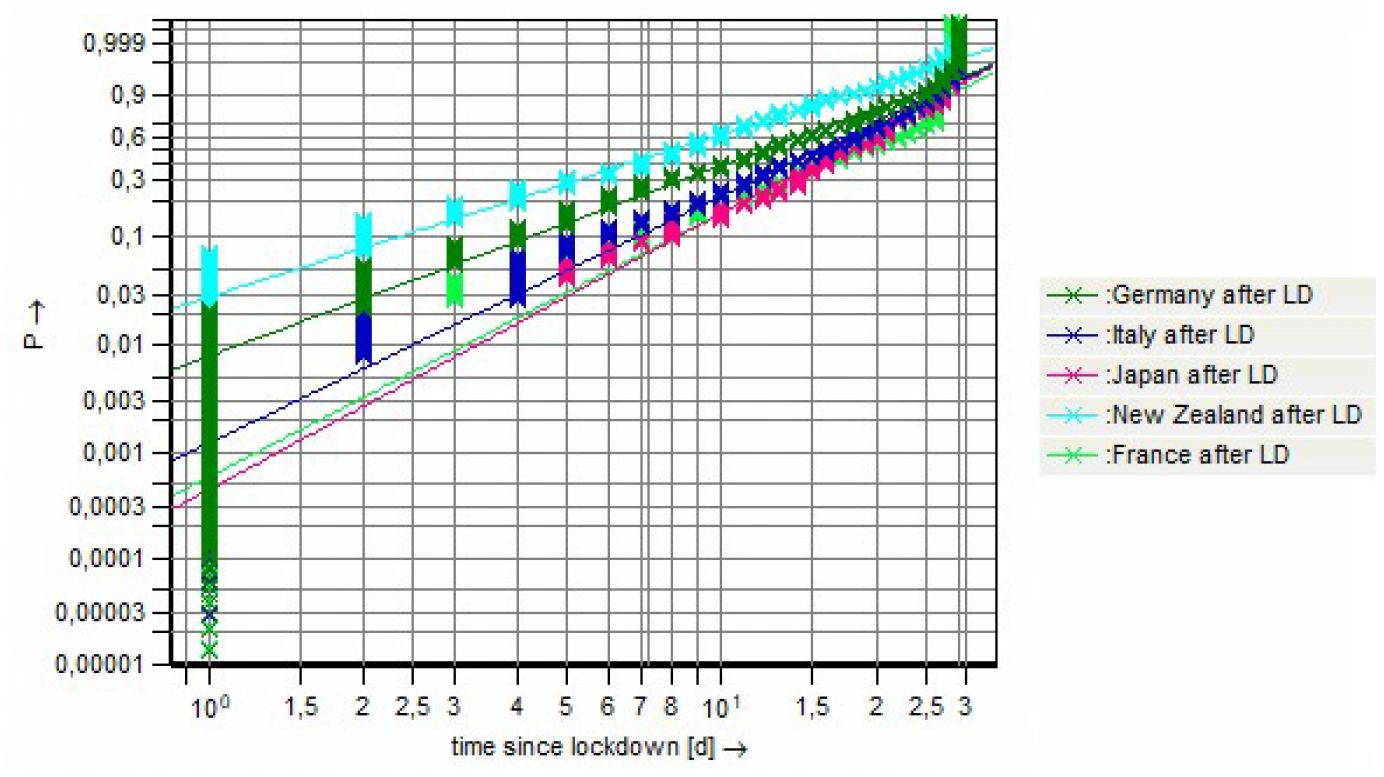
Weibull distribution models, representing the infection development, confirmed cases, time span 28 days from the lockdown related to the analyzed countries (log-log-scale).

The estimated Weibull parameters are shown in Table 3.

**Table 3:** Weibull model parameters since lockdown (confirmed cases, approx. 28 days) related to analyzed countries. Confidence level γ = 0.95.

| Country | cases | T [d] | Shape b [confidence belt] |
| --- | --- | --- | --- |
| Germany | 122,971 | 15 | $1.78 \leq 1.79 \leq 1.80$ |
| Italy | 126,415 | 18 | $2.31 \leq 2.32 \leq 2.33$ |
| Japan | 11,765 | 20 | $2.56 \leq 2.60 \leq 2.64$ |
| New Zealand | 1,173 | 10 | $1.46 \leq 1.53 \leq 1.60$ |
| France | 121,605 | 20 | $2.47 \leq 2.48 \leq 2.50$ |

First, the reduction of the shape parameter within the comparison before and after lockdown (cf. Table 2 and 3) is clearly visible for all analyzed countries. The change is significant due to comparison of confidence intervals of the shape parameters (before/after lockdown). The spreading speed (gradient) is significantly reduced, the biggest change is observed in Germany. The smallest spreading speed after lock-down can be seen in New Zealand. There, strict border closures resulted in a halt to the spreading of the virus; the gradient (shape parameter) is close to a random failure rate characteristic (shape parameter b ∼ 1). Besides, the low population density (18.6/km^2^) can be a reason for this low spreading speed. Japan shows the highest shape parameter (spreading speed) after lockdown in comparison to the other countries, maybe explainable by the highest population density (334/km^2^). Though these differences in population density (uncertainty factor), the characteristics of the lock- down (soft, hard, etc.) in the different countries can be interpreted as the reason for the different Weibull distribution model fit results.

### 6.3 Second wave detection

To detect the beginning of the second wave, a trend test is done. Therefore, since 1 July, the daily confirmed cases are analyzed. At this time, the spreading of the first wave had decreased and there were few new case numbers.

A one-sided trend test (upward trend) according to Cox and Stuart (1955) is performed with 14 data points and a significance level α of 5%, cf. Eq. 2. The tested hypotheses are as follows:

- Null hypothesis: There is no upward trend.
- Alternative hypothesis: There is an upward trend (second wave).

The sample size of 14 days is chosen to mitigate outliners and data falsifications like the weekend impact. If the p value of the test is smaller than α, the date of the first data point in the tested data set is detected as the beginning of the second wave. Otherwise, the sample is moved one day to the right and the test is performed again. The Cox and Stuart significance test was performed 71 times in this way on the example of Germany; cf. Fig. 3. The results are the p values as black points plotted on the right ordinate. As a comparison, the daily confirmed cases are deducted on the left ordinate. The corresponding dates are assigned to these values. Additionally the significance is shown as a horizontal red line.

**Fig. 3:**
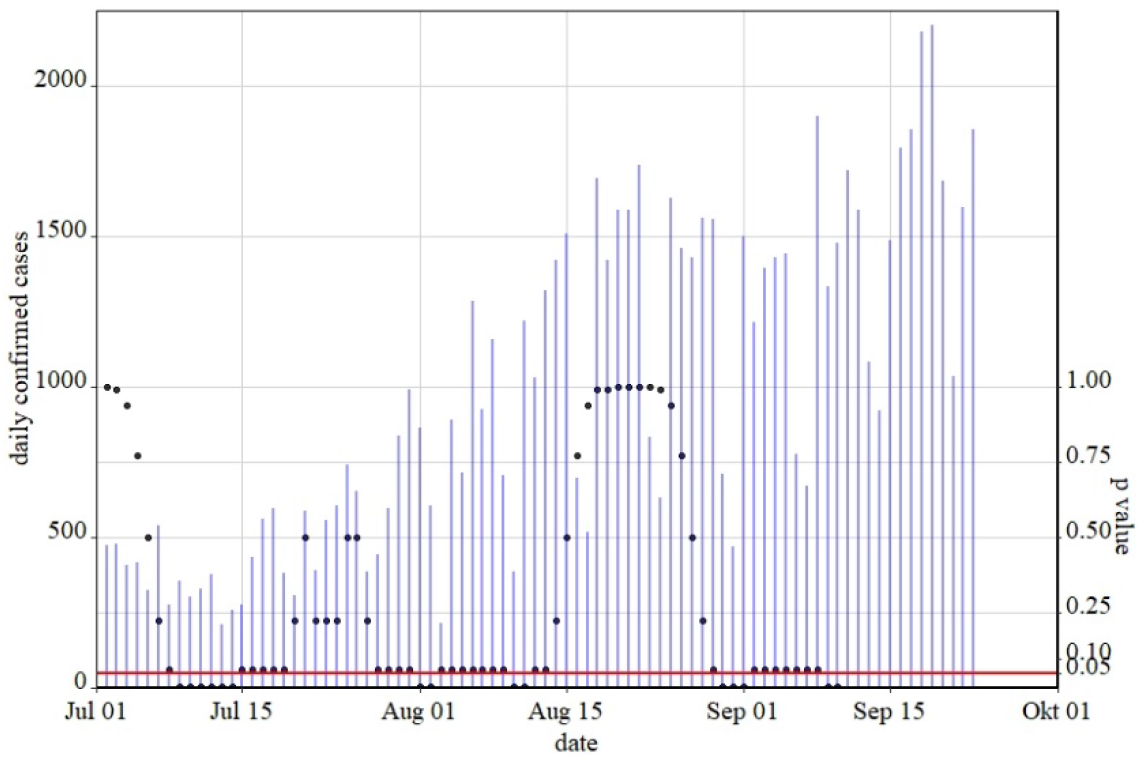
Second wave detection with Cox-Stuart trend test, p values (black points), alpha=0.05 (red line), daily confirmed cases Germany since 01 July (blue). Data inventory 09/22/2020.

All points placed under the red line represent those tests, which result in an upward trend. According to the described approach, the first point under the red line represents the beginning of the second wave; here the p value is below the significance level, so the null hypothesis is rejected and the alternative hypothesis of an upward trend is assumed. Based on these analyzing procedure, in Germany the second wave started on 8 July 2020. In comparison to the daily confirmed cases this date seems plausible for the beginning of a second wave, from then on an increase in the number of cases can be seen.

The results of the trend tests for the other countries and thus the respective beginnings of the second wave are documented in Table 4.

**Table 4:**
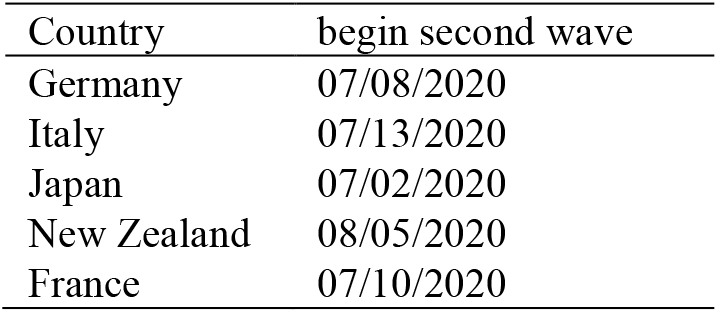
Detected begin second wave per country, based on application Cox and Stuart significance test.

A European wide second wave can be recognized; the beginnings of the second waves of Germany, Italy and France are in a similar period. The second wave in Japan started earlier, while the New Zealand second wave began about one month later.

### 6.4 Infection (confirmed cases) second wave

The spreading of the early second wave is analyzed with a Weibull model fit. Therefore, a period of about 50 days after the beginning of the second wave is considered for the analyzed countries: Germany, Italy, Japan, New Zealand and France. This time span allows a comparison with the shape parameter (spreading speed) of the first wave, the spreading until lockdown, due to the same amount of analyzed days. Fig. 4 shows the Weibull distribution models of the second wave. the shape parameter is the smallest, and so the second wave is not very pronounced there, which is consistent with the low number of cases.

**Fig. 4:**
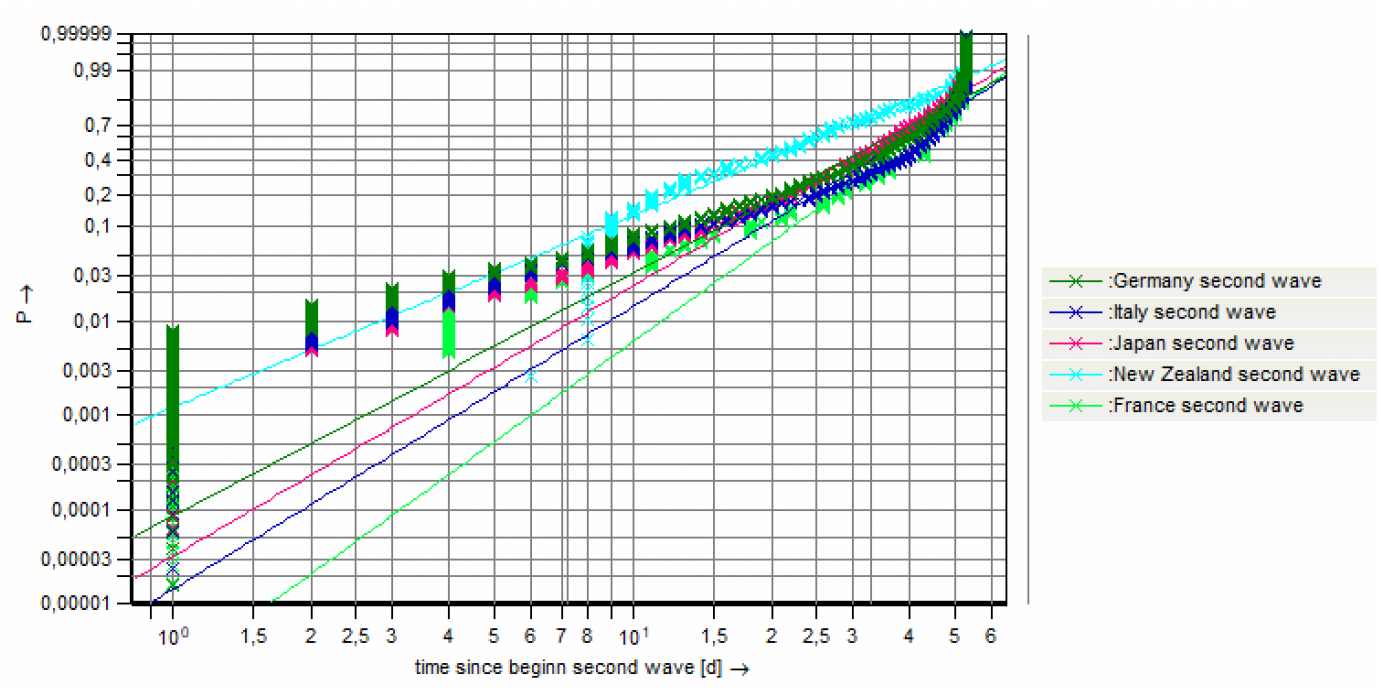
Weibull distribution models, representing the infection development, confirmed cases, time span 50 days from the second wave related to the analyzed countries (log-log-scale).

The estimated Weibull parameters are shown in Table 5.

**Table 5:** Weibull model parameters since begin second wave (confirmed cases, 50 days) related to analyzed countries. Confidence level γ = 0.95.

| Country | cases | T [d] | Shape b [confidence belt] |
| --- | --- | --- | --- |
| Germany | 44,492 | 37 | $2.58 \leq 2.60 \leq 2.61$ |
| Italy | 29,851 | 41 | $2.99 \leq 3.01 \leq 3.04$ |
| Japan | 43,820 | 37 | $2.86 \leq 2.88 \leq 2.90$ |
| New Zealand | 264 | 27 | $1.85 \leq 2.05 \leq 2.25$ |
| France | 105,860 | 42 | $3.52 \leq 3.54 \leq 3.56$ |

France has the highest spreading speed, which corresponds to the high number of cases in a short time (hard second wave). The curve of New Zealand is the flattest, the shape parameter is the smallest, and so the second wave is not very pronounced there, which is consistent with the low number of cases.

When comparing the shape parameter with that of the first wave, it is noticeable that the early second wave shows a spreading spread on a lower (moderate) level. Rather, the propagation speed is in the range of the course after the lockdown. By comparing the confidence intervals of the gradient (shape parameter b), it becomes clear that the spreading speed of the early second wave is significantly higher than that of the time after the lockdown.

The spreading speed of the early second wave is lower than that of the first wave and slightly higher than the one of the time after the lockdown. It has to be considered, that the data inventory of this paper is the 09/22/2020, so further developments of the second wave are not analyzed.

## 7 Comparison of the COVID-19 spreading behavior with other infectious diseases

To see the COVID-19 spreading in a greater context, it is compared with the spreading of influenza and measles in Germany. The seasons 2014/15 until 2016/17 are chosen as comparable seasons with average case numbers and process. The first 56 days since the first case are analyzed for each season in a first step. Then, a 3- year average is estimated and the spreading is compared with COVID-19 before and after lockdown.

### 7.1 Influenza and measles

To gain the knowledge regarding the spreading speed of typical infectious diseases, Weibull distribution models are fitted to in each case three seasons of influenza and measles. To create a comparability with the COVID-19 spreading, the first 56 days (8 weeks) are analyzed. Since the data from RKI (2020) is given on a weekly base, the analyzing of the ranked data is based on a transformation (1 week corresponds to 7 days). Fig. 5 shows the resulted Weibull distribution models, the Weibull parameters are listed in Table 6.

**Table 6:** Weibull model parameters since index case (56 days) related to analyzed diseases and seasons. Confidence level γ = 0.95.

| Disease | Season | cases | T [d] | Shape b [confidence belt] |
| --- | --- | --- | --- | --- |
| Influenza | 14/15 | 116 | 43 | $2.52 \leq 2.93 \leq 3.39$ |
| | 15/16 | 266 | 47 | $3.18 \leq 3.52 \leq 3.87$ |
| | 16/17 | 510 | 45 | $2.59 \leq 2.78 \leq 2.98$ |
| Measles | 14/15 | 72 | 38 | $1.91 \leq 2.32 \leq 2.78$ |
| | 15/16 | 22 | 30 | $1.38 \leq 2.01 \leq 2.77$ |
| | 16/17 | 128 | 40 | $2.48 \leq 2.87 \leq 3.29$ |

**Fig. 5:**
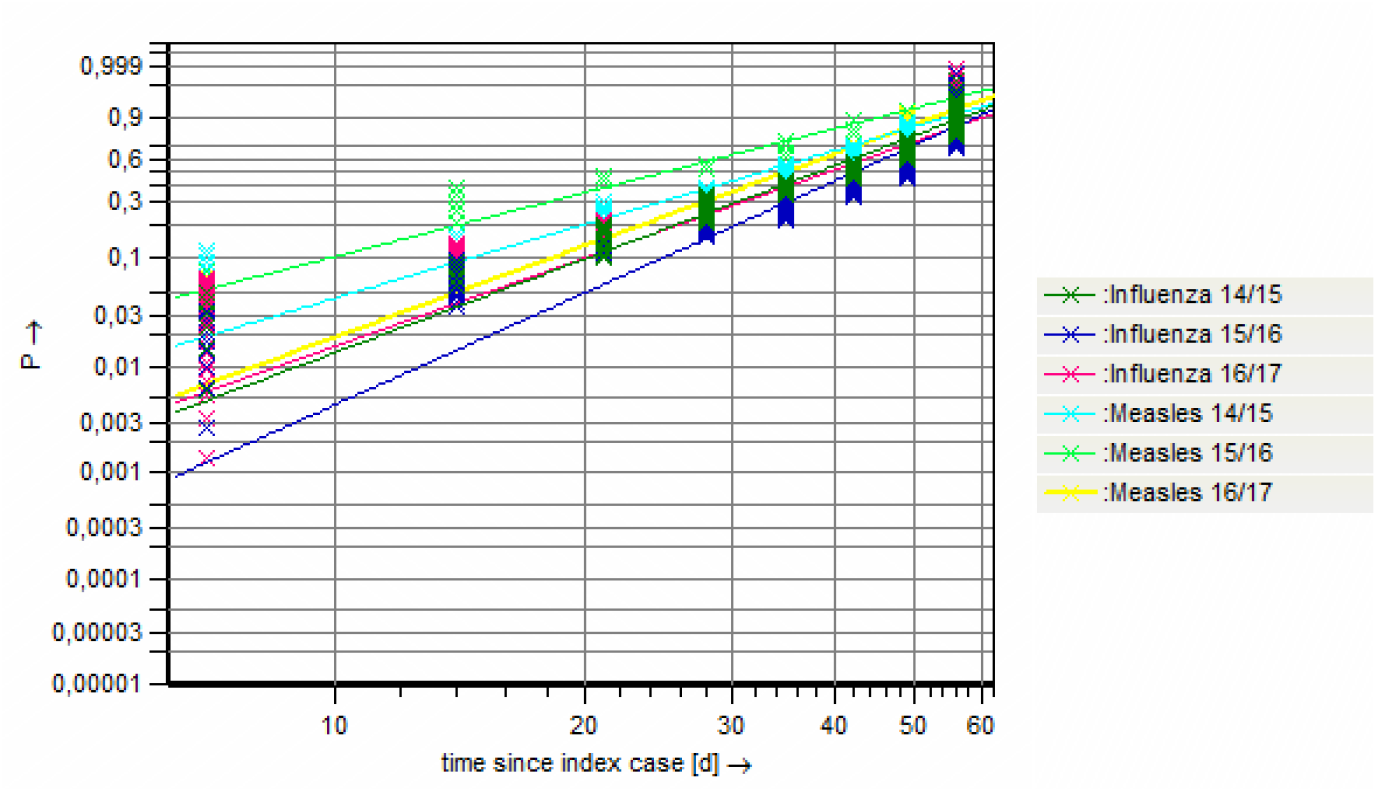
Weibull distribution models, representing the infection development, influenza and measles, time span 56 days from the index case related to the analyzed seasons (log-log-scale).

There are more influenza cases than measles cases, the parameters variate between the seasons and the disease. The spreading speed (shape parameter) of measles is slightly slower than that of influenza. Since the confidence intervals of the two diseases overlap, this difference is not significant. In addition, the spreading speeds within the diseases slightly, not significant, differ. Therefore, the summarization in an average of the seasons (3-year average) is legitimate, as done in the next step.

### 7.2 Comparison: COVID-19 versus Influenza versus Measles

On the base of the known infection cases, a 3-year average is developed for influenza and measles. Weibull distribution models are fitted for these averages and the COVID-19 spreading before and after lockdown on the example of Germany.

The comparison of COVID-19 / influenza / measles models is shown in Fig. 6 and the corresponding parameters are documented in Table 7. It is clearly visible that the COVID-19 spreading differs from the spreading of other infectious diseases like influenza and measles. The spreading speed (shape parameter) of COVID-19 before lockdown is significant larger (influenza: factor ∼6.6; measles: factor ∼6.3) than that of the 3-year averages of influenza and measles. This shows that the extreme progress of COVID-19 pandemic in comparison to influenza and measles. The spreading behavior of COVID-19 is not comparable with “normal” infectious diseases.

**Table 7:** Weibull model parameters since index case (about 50 days) and since lockdown LD; 28 days) related to analyzed diseases. Database: Germany. Confidence level γ = 0.95.

| Disease | cases | T [d] | Shape b<br>[confidence belt] |
| --- | --- | --- | --- |
| COVID-19 before LD | 22,213 | 53 | $19.61 \leq 19.82 \leq 20.03$ |
| COVID-19 after LD | 122,971 | 15 | $1.78 \leq 1.79 \leq 1.80$ |
| Influenza 3-year average | 296 | 45 | $2.73 \leq 3.00 \leq 3.28$ |
| Measles 3-year-average | 75 | 43 | $2.58 \leq 3.13 \leq 3.74$ |

**Fig. 6:**
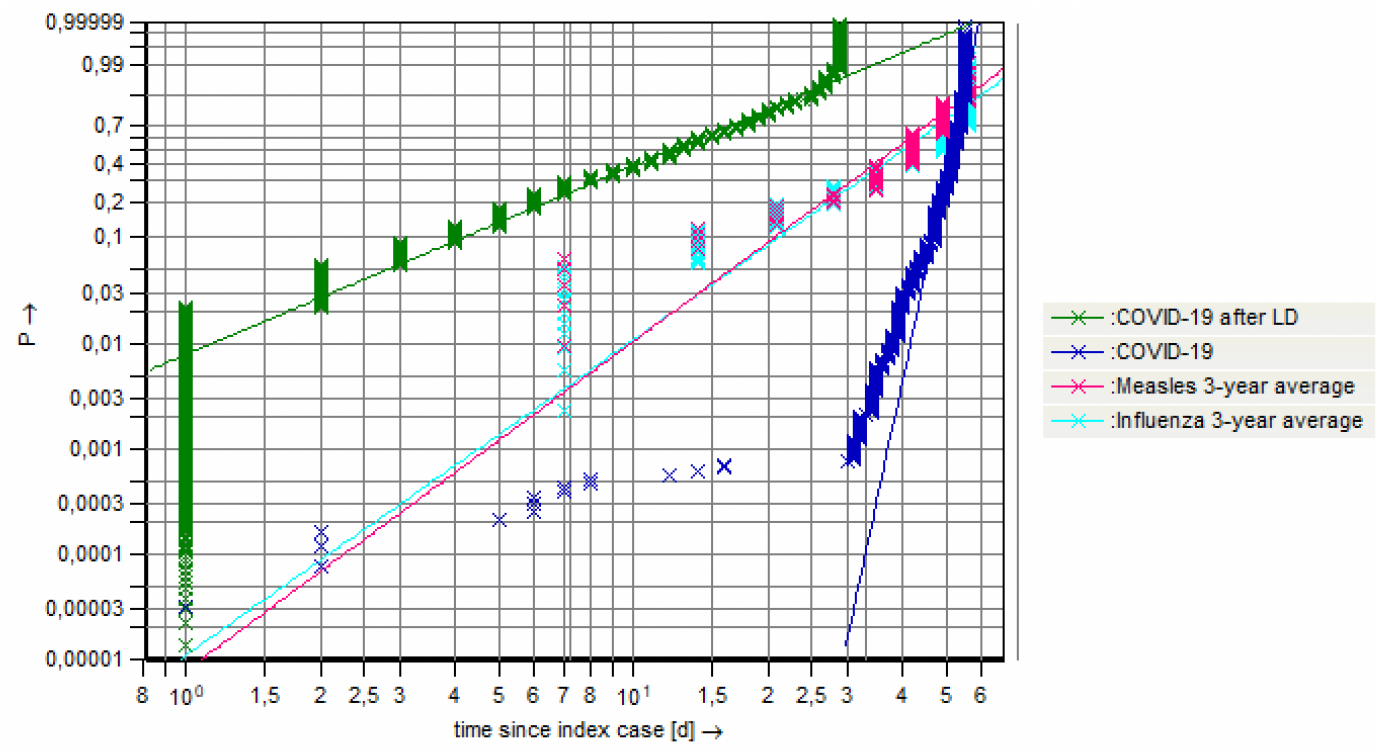
Weibull distribution models, representing the infection development, comparison of COVID-19 before and after lockdown with 3-year average of measles and influenza 2013/14 -2015/16, time span about 50 days (log-log-scale). Database: Germany.

In the 3-year average, no significant difference can be seen between the spreading speed of measles and influenza. Obviously, the on hard measures of the lock- down (e.g. restrictions on contact, distance regulations) lead to the level of a “normal” infectious diseases (influenza and measles) spreading behavior. Without any measures, the spreading speed (gradient) of COVID-19 is higher by factor ∼6.6 compared to influenza respectively by factor ∼6.3 compared to measles.

Note: The spreading of influenza and measles is also affected by basic immunity caused by available vaccines, which do not exist for COVID-19 disease.

## 8 Summary

The application of statistical methods of the reliability engineering enabled a detailed analysis of the occurrence of infection. Using Weibull distribution models, the spreading behavior can be evaluated. The main part of the data analytics was the application of a model used for damage cases within reliability engineering analytics describing the cases of a pandemic. The shape parameter b as the gradient of the Weibull distribution model (log-log-scale) is interpreted as the spreading speed. When analyzing data from different countries, uncertainty factors and data quality must be taken in account: Differences in the countries like test strategies or reporting systems, definitions of cases or external influences like seasonality effect the database. This was handled by analyzing ranked data and countries with similar industrialization standard.

By comparing the shape parameters and their confidence intervals from the time before and after the lockdown, it got clear that the lockdown could significantly reduce the COVID-19 spreading speed in all analyzed countries.

The Cox and Stuart trend test detected the beginning of the second wave depending from the analyzed country in July or August 2020. Thereby a European wide second wave could be determined, the detected starting points are within short time range. In terms of spreading speed, it could be noticed that the early second wave is much more moderate than the first wave in terms of spreading. In comparison to the time after the lockdown, the spreading speed is higher. With the data inventory of the 09/22/2020, further developments cases were not analyzed and the impact of autumn respectively winter season is not considered. Considering these seasonal effect, a mixed distribution – with different increasing gradients - can be expected. Further research studies will focus on a longterm comparison of first and second COVID-19 wave.

To see the COVID-19 spreading in a greater context, it was compared with the spreading of influenza and measles on the example of Germany. Thereby, the Weibull distribution model was fitted to the first time of the season to enable a comparison with the COVID-19 spreading. It got clearly visible that the COVID- 19 spreading differs from the spreading of other infectious diseases. The spreading speed (shape parameter) of COVID-19 before lockdown is significant higher with factor ∼6.6 in comparison to the 3-year averages of influenza, and higher with factor ∼6.3 in comparison to measles. This shows that the extreme progress – the spreading speed – of COVID-19, it is not comparable with “normal” infectious diseases (influenza/measles). Only the COVID-19 time period under the strong impact of the lockdown measures, shows a spreading speed on the level of influenza or measles.

## Data Availability

JHU - Johns Hopkins University - COVID-19 Dashboard
RKI - Robert Koch Institute

https://gisanddata.maps.arcgis.com/apps/opsdashboard/index.html#/bda7594740fd40299423467b48e9ecf6

https://survstat.rki.de

